# An Interpretable Deep Learning Model for Early Prediction of Sepsis in the Emergency Department

**DOI:** 10.1101/2020.09.21.20198895

**Authors:** Dongdong Zhang, Changchang Yin, Katherine M. Hunold, Xiaoqian Jiang, Jeffrey M. Caterino, Ping Zhang

**Author notes:** Equal contributor (Dongdong Zhang, Changchang Yin).

## Abstract

**Background:** Sepsis, a life-threatening illness caused by the body’s response to an infection, is the leading cause of death worldwide and has become a global epidemiological burden. Early prediction of sepsis increases the likelihood of survival for septic patients.

**Methods:** The 2019 DII National Data Science Challenge enabled participating teams to develop models for early prediction of sepsis onset with de-identified electronic health records of over 100,000 unique patients. Our task is to predict sepsis onset 4 hours before its diagnosis using basic administrative and demographics, time-series vital, lab, nutrition as features. An LSTM-based model with event embedding and time encoding is proposed to model time-series prediction. We utilized the attention mechanism and global max pooling techniques to enable interpretation for the proposed deep learning model.

**Results:** We evaluated the performance of the proposed model on 2 use cases of sepsis onset prediction which achieved AUC scores of 0.940 and 0.845, respectively. Our team, *BuckeyeAI* achieved an average AUC of 0.892 and the official rank is #2 out of 30 participants.

**Conclusions:** Our model outperformed collapsed models (i.e., logistic regression, random forest, and LightGBM). The proposed LSTM-based model handles irregular time intervals by incorporating time encoding and is interpretable thanks to the attention mechanism and global max pooling techniques.

**Availability:** The code for this paper is available at: https://github.com/yinchangchang/DII-Challenge.

## Background

Sepsis, a life-threatening illness caused by the body’s response to an infection, is the leading cause of death worldwide and has become a global epidemiological burden. Sepsis occurs at all ages and increases mortality rate. In the United States, for example, over 1.7 million adults develop sepsis and nearly 270,000 patients die as a result of sepsis each year [1]. Besides, sepsis is the costliest among all disease states and accounts for $24 billion of U.S. hospital costs in 2013 [2].

Sepsis-2 is diagnosed as the presence of proven or suspected infection together with 2 or more systemic inflammatory response syndrome (SIRS) criteria [3]. Without timely and adequate treatment, sepsis can progress to severe sepsis and septic shock, which lead to higher mortality. Several studies suggest that early prediction of sepsis enables early treatment and significantly improves patient outcomes [4, 5].

Electronic Health Records (EHRs) are longitudinal electronic records of patients’ health information and the percent of non-Federal acute care hospitals with the adoption of at least a Basic EHR system increased from 9.4% to 83.8% over the 7 years between 2008 and 2015 in the United States [6]. The broad adoption of electronic health records (EHRs) provides great opportunities for conducting health care research. With recent advances and success, machine learning methods have shown great potential in unlocking insights from EHRs. Various methods have been developed for accurate sepsis prediction [7, 8]. Faisal [9] developed a logistic regression model (CARS) to predict the risk of sepsis using a patient’s firstly recorded vital signs and blood test results, which are usually available within a few hours of emergency admission. Horng [10] constructed a machine learning model using a linear support vector machine (SVM) and demonstrated the incremental benefit of using free text data in addition to vital signs and demographic data for sepsis clinical decision support at the emergency department. Lyra [11] used an optimized random forest to predict sepsis for imbalanced clinical data from intensive care units in the PhysioNet Computing in Cardiology Challenge 2019 [8]. Mao [12] validated a machine learning algorithm with gradient boosting trees, *InSight*, which used only 6 vital signs for the prediction of sepsis, severe sepsis, and septic shock and showed that *InSight* outperformed existing sepsis scoring systems. Using 65 features from a combination of EHR and high-frequency physiological data, Nemati [13] developed and validated an interpretable machine learning model based on modified Weibull-Cox proportional hazards algorithm for making an accurate and interpretable prediction of sepsis.

Recently, deep learning methods have achieved promising results and shown unprecedented potential in many areas [14] such as computer vision [15], speech recognition [16], and natural language processing [17]. Deep learning models automatically learn the data representation with improved performance and do not require conventional feature extraction steps. Recurrent neural networks (RNNs) and convolutional neural networks (CNNs) are commonly used network architectures in modeling multivariate series prediction [18, 19, 20]. van [21] built a convolutional neural network model to classify septic and non-septic patients and the CNN-based model outperformed a multi-layer perceptron model for various data collection frequencies. Kam [22] proposed a sepsis detection model with long short term memory (LSTM) and their model showed better performance than *InSight* and superior capability for sequential patterns. Fagerström [23] developed an improved algorithm, LiSep LSTM, and demonstrated the benefit of using an LSTM network as opposed to the Cox proportional hazards model for early prediction of septic shock. Lauritsen [24] proposed a deep learning model based on a combination of a convolutional neural network and a long short-term memory network (CNN-LSTM) that could learn characteristics of the key factors and interactions from the raw event sequence data and outperformed baseline models.

Common signs and symptoms of sepsis, such as fever, chills, rapid respiratory, and high heart rate, are the same as in other conditions, making sepsis hard to diagnosis in its early stages. The DII (Discover, Innovative, Impact) challenge is a national data science challenge established to advance human health through machine learning. The goal of the 2019 DII challenge is the early prediction of sepsis using a patient’s demographic and physiological data. The 2019 DII challenge provided an opportunity for researchers to develop machine learning and deep learning methods to computationally detect sepsis 4 hours ahead before onset.

Our team, *BuckeyeAI*, participated in the 2019 DII challenge and ranked #2 out of 30 teams on the early prediction of sepsis onset task with an average AUC score of 0.892. In this paper, we presented our methods, results, and analyses. To summarize, the contributions are:

- We present benchmark results of the sepsis onset prediction task. We show that our model outperforms baseline machine learning models.
- We propose an LSTM-based model for sepsis onset prediction which handles irregular time intervals with time encodings.
- We leverage the attention mechanism and global max pooling techniques to help interpret our model.

## Method

### Dataset description

#### Definition of sepsis

The goal of 2019 DII challenge is early prediction of sepsis with demographic and physiological data provided. In this challenge, definition of sepsis2, presence of proven or suspected infection together with 2 or more SIRS criteria, is used as the gold standard. The SIRS criteria are defined as:

- heart rate *>* 90 beats/min
- body temperature *>* 38^°^C or *<* 36^°^C
- respiratory rate *>* 20 breaths/min or PaCO2 *<* 32 mm Hg
- white cell count *>* 12×10^9^ cells/L or *<* 4 ×10^9^ cells/L

#### Cohort preparation and statistics

The challenge data are extracted from the Cerner Health Facts database. Cerner Health Facts is a database that comprises de-identified EHR data from over 600 participating Cerner client hospitals and clinics in the United States and represents over 106 million unique patients. With this longitudinal, relational database—reflecting data from 2000-2016—researchers can analyze detailed sets of de-identified clinical data at the patient level. Types of data available include demographics, encounters, diagnoses, procedures, lab results, medication orders,medication administration, vital signs, microbiology, surgical cases, other clinical observations, and health systems attributes. Sepsis2 definition is used to define the ground truth. Patients who are *<* 18 years old or do not have enough observation data are excluded. The whole data preparation pipeline diagram is shown in Figure 1. The label statistics and characteristics of the final cohort are provided in Table 1.

**Table 1.** Label statistics and characteristics of the final cohort.

|  |  | Sepsis2 patient | Sepsis2 control |
| --- | --- | --- | --- |
| Total |  | 52,802 (29.5%) | 126,041 (70.5%) |
| Gender | Female | 25,936 (49.1%) | 65,523 (52.0%) |
|  | Male | 26,866 (50.9%) | 60,518 (48.0%) |
| Race | African American | 11,084 (21.0%) | 20,556 (16.3%) |
|  | Asian | 1,085 (2.1%) | 1,627 (1.3%) |
|  | Caucasian | 35,059 (66.4%) | 95,657 (75.9%) |
|  | Others/unknown | 5,574 (10.5%) | 8,201 (6.5%) |
| Age | 18-20 | 1,602 (3.0%) | 1,776 (1.4%) |
|  | 20-40 | 8,100 (15.3%) | 15,288 (12.1%) |
|  | 40-60 | 15,654 (29.6%) | 34,295 (27.2%) |
|  | 60-80 | 20,241 (38.3%) | 51,914 (41.2%) |
|  | 80-100 | 7,205 (13.6%) | 22,768 (18.1%) |

**Figure 1.**
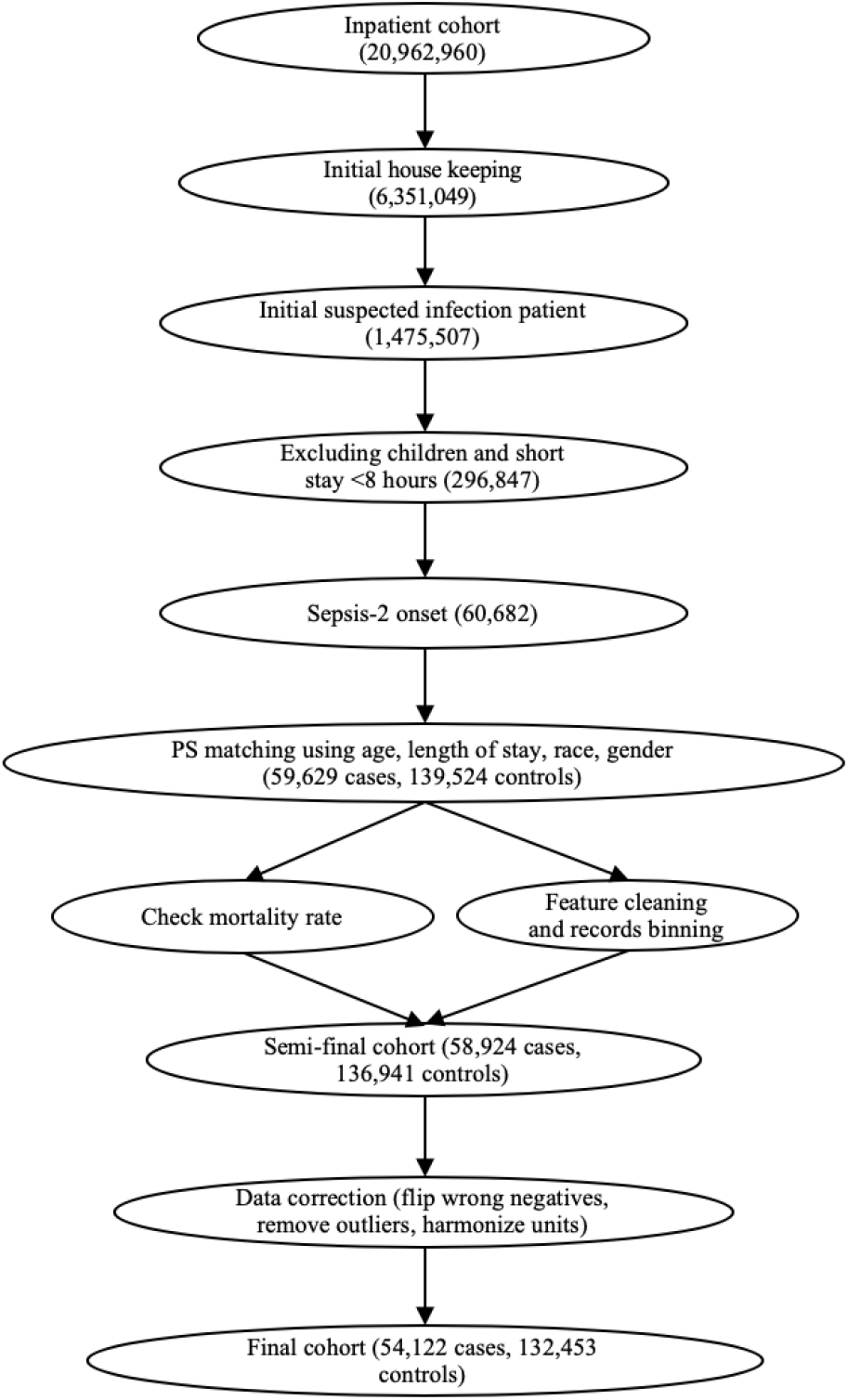
Inclusion and exclusion diagram of DII challenge data preparation pipeline. After filtering and correction, the final cohort has a sepsis prevalence of 29.0%.

### Predictive tasks and evaluation metric

In this challenge, we aim to predict sepsis 4 hours before onset for hospitalized adult patients. There are 2 use cases as demonstrated in Figure 2.

**Figure 2.**
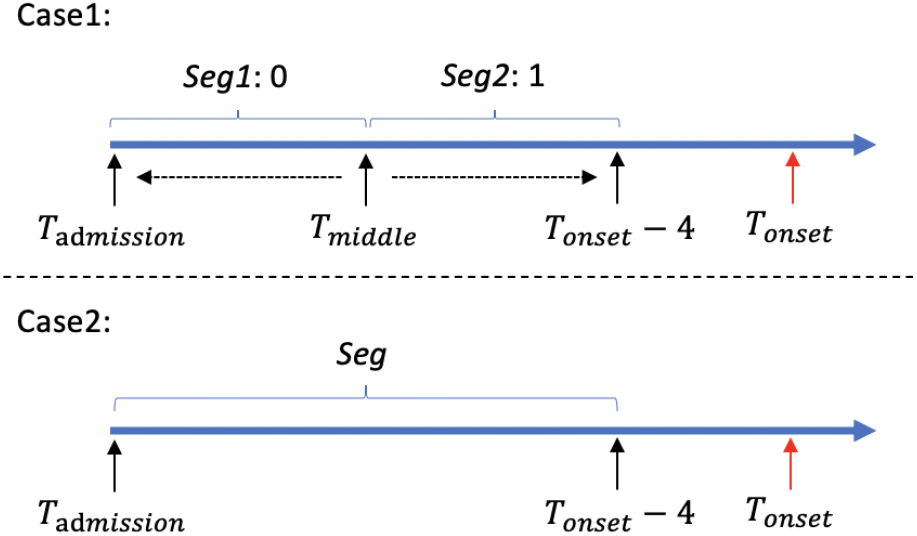
Two use cases of sepsis onset prediction 4 hours before it occurs.

*Case 1* In this case, patients are sampled from septic patients. By randomly splitting patient records into 2 segments at a roughly middle point, segment close to sepsis onset (= 4 hours) is labeled as 1, another segment (*>* 4 hours before sepsis onset) is labeled 0. Given patient records either from *T*_*admission*_ to *T*_*middle*_ or from *T*_*middle*_ to *T*_*onset*_ −4, our model is required to distinguish this 2 kinds of records.

*Case 2* In this case, case and control segments are from different patients who have sepsis onset in the next 4 hours, as well as those who do not have sepsis. Given patient records from *T*_*admission*_ to *T*_*onset*_ −4, we are going to predict whether sepsis occurs in the following 4 hours.

To evaluate the performance and discrimination of binary classifier, for each use case, the area under the receiver operating characteristic curve (AUC) is used as the evaluation metric. The arithmetic average of AUC scores of 2 use cases is used for final performance comparison.

### Neural network architecture

The proposed neural network architecture is shown in

This model is inspired by [25]. Although we focus on the early prediction of sepsis onset in this challenge, our proposed model is general and can be applied to other multivariate time-series prediction tasks, such as mortality prediction for septic patients.

#### Event embeddings

For each temporal feature, we sort the values from low to high and use the order to replace the original values. Then, we divide the orders into 10 groups (i.e., 0.0-0.1, 0.1-0.2, …, 0.9-1.0) and then each event is embedded into a 512-d vector.

#### Time encodings

When modeling time-series EHR data, most existing LSTM-based models only consider the relative order of events. However, these methods typically ignore the irregular time intervals between neighboring events. Similar to position encodings in Transformer [26], we infuse time information using “time encodings”. Time encodings are sent to LSTM together with value embeddings. We compute each event’s relative time to the criterion operation date and the time interval relative to the last event. Then, we use sine and cosine functions of the different time intervals to represent the time encoding for the *t*_*th*_ event:

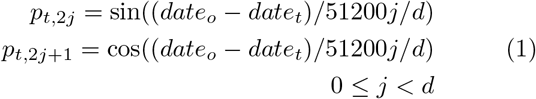

where *data*_*o*_ denotes the criterion operation date, *data*_*t*_ denotes the *t*_*th*_ event’s date, and *p*_*t*_ ∈ *R*_2*d*_ denotes the time encoding vector, and *j* is the dimension of EHRs event embeddings. Then both the event embeddings and time encodings are input to LSTM.

To better align patient records at their last recorded medical event, the time of each event is mapped from [0, *T*_*lastevent*_] to [−*T*_*lastevent*_, 0].

#### LSTM and attention mechanism

RNNs are popular and suitable for sequential EHR data modeling. Given medical event embedding and time encoding vectors, we build our model based on LSTM [27] for its ability to recall long term information. The LSTM model can be described as follows:

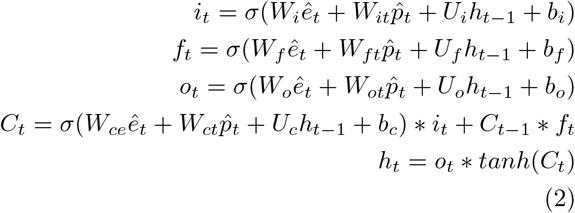

where *σ* is the sigmoid function, *t* denotes the *t*_*th*_ step of LSTM, and *C*_*t*_ is the corresponding cell state, and *h*_*t*_is the output vector. *ê*_*t*_ is the input event embedding, 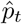 is the input time enconding. *W*_*i*_, *W*_*f*_, *W*_*o*_, *W*_*ce*_ ∈ *R*^*k*×*d*^, *W*_*it*_, *W*_*ft*_, *W*_*ot*_, *W*_*ct*_ ∈ *R*^*k*×2*d*^, *U*_*i*_, *U*_*f*_, *U*_*o*_, *U*_*c*_ ∈ *R*^*k*×*d*^, *b*_*i*_, *b*_*f*_, *b*_*o*_, *b*_*c*_ ∈ *R*^*k*^ are learnable parameters. Attention mechanism is used to automatically identify influential clinical features.

#### Global max pooling

RNN-based models are sometimes inefficient due to their long-term dependency. When the input sequence is too long, it is easy for the models to forget the earlier data. Therefore, we adopt a global pooling operation to shorten the distance between the earlier inputs and the final outputs. As is shown in Figure 3, all the outputs of the LSTM are concatenated, and then a global pooling operation is followed. The output *o*_*g*_ is fed through the fully connected layer to produce the clinical risk of patient *i*, which is defined as:

**Figure 3.**
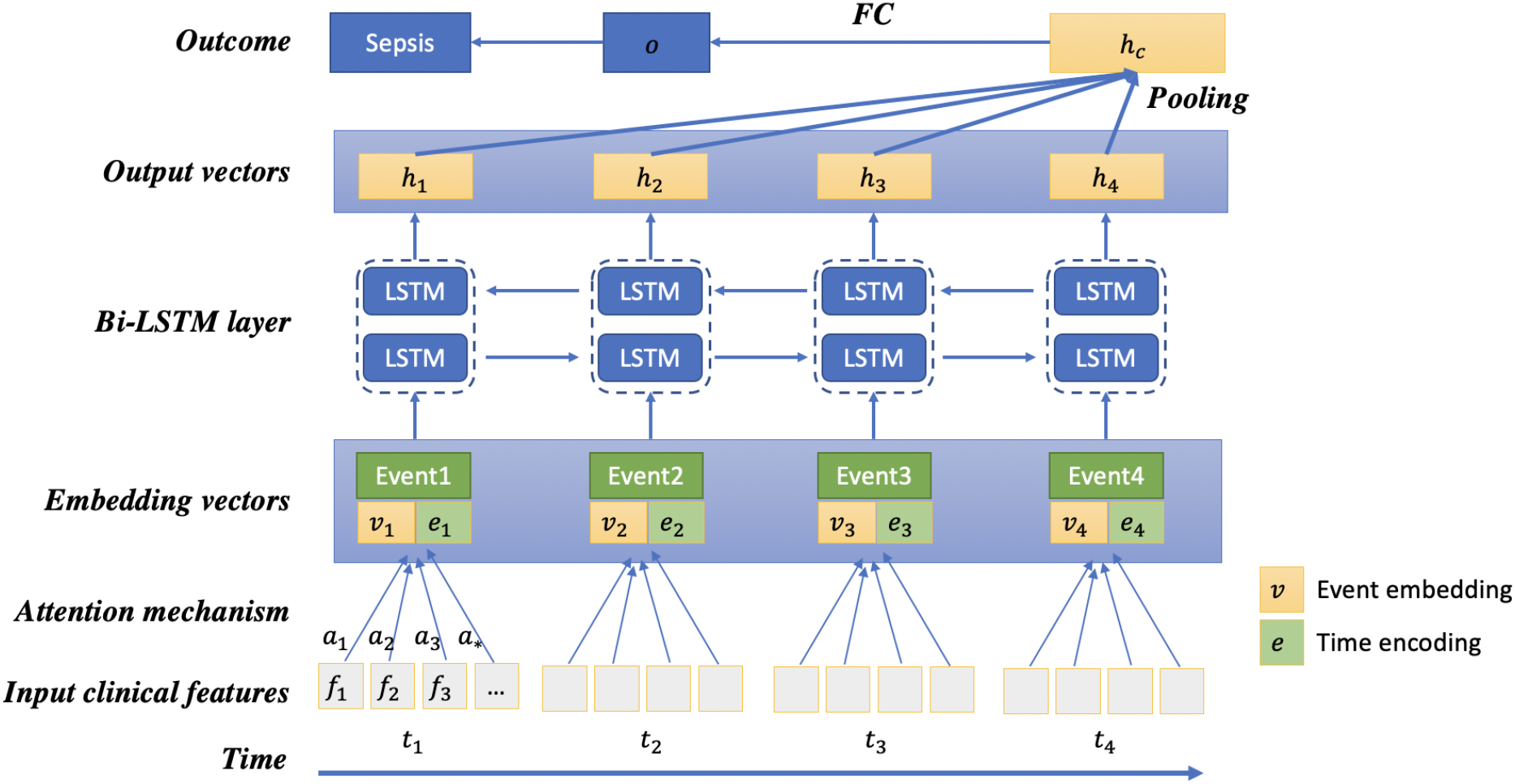
Architecture of proposed LSTM-based model. The concatenation of the medical event embedding vectors (*v*_1_, *v*_2_,…, *v*_*n*_) and the corresponding time encoding vectors (*e*_1_, *e*_2_,…, *e*_*n*_) are inputs to the model. For each input event, the Bi-LSTM model generates 2 output vectors. All the output vectors are concatenated and then a global max pooling operation is followed to produce the patient representation vector. Finally, a fully connected layer and the sigmoid function are used to predict the probability of sepsis onset in the next 4 hours.

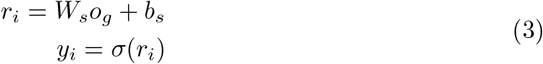

where *W*_*s*_ ∈ *R*^*k*^ and *b*_*s*_ ∈ *R* are the learnable parameters, *y*_*i*_ denotes the predicted probability for sepsis onset. Because of the shortened distance between the inputs and the outputs, the pooling operation makes it more efficient to propagate the gradients. Besides, the global pooling operation is useful to compute the contribution rates of the outputs and their corresponding input medical events.

#### Objective function

For binary classification, the objective function is defined as the binary cross entropy loss between ground truth *y*^*^ and predicted probability *y*:

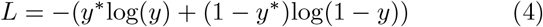

#### Interpretability

Interpretability is very important for machine learning models of clinical applications. The global pooling operation leveraged in our architecture can associate the contribution of each input medical event to the final clinical outcome, paving the way for interpretable clinical risk predictions.

In Figure 3, given the output vectors, the global max pooling operation is followed and produced the final patient feature vector *h*_*c*_, which is used to predict risk of sepsis onset. We can track the output vectors which constitutes specific element of *h*_*c*_. After the fully connected layer, we can calculate every dimension’s contribution rate. For a case patient, the contribution rate of output vector *h*_*t*_ for the *t*_*th*_ input event is is calculated as:

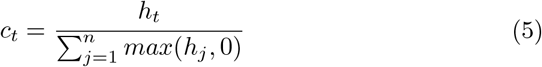

In order to illustrate the interpretability of our model clearly, we display 2 input events and 4 corresponding 6-dimensional output vectors (*h*_1_, *h*_2_, *h*_3_, *h*_4_) in Figure 4.Given patient feature vector (*h*_*c*_) and fully connected parameters (*W*_*s*_, *b*_*s*_), the output risk is computed (*r*_*i*_ = *W*_*s*_*h*_*c*_ + *b*_*s*_). The first dimension’s contribution risk is 0.21 and the contribution rate is 9.1%, which comes from the fourth output vector *h*_4_. Similarly, the fifth dimension’s contribution rate also comes from *h*_4._ Thus, the contribution rate of the fifth vector *h*_4_ is computed by summing the two contribution rates. Then, we compute the contribution rate of the input event *e*_2_ by summing the contribution rates of *h*_3_ and *h*_4_, *c*_2_ = 36.9%. For feature *j* in event *i*, we can compute its contribution rate with attention weight as:

**Figure 4.**
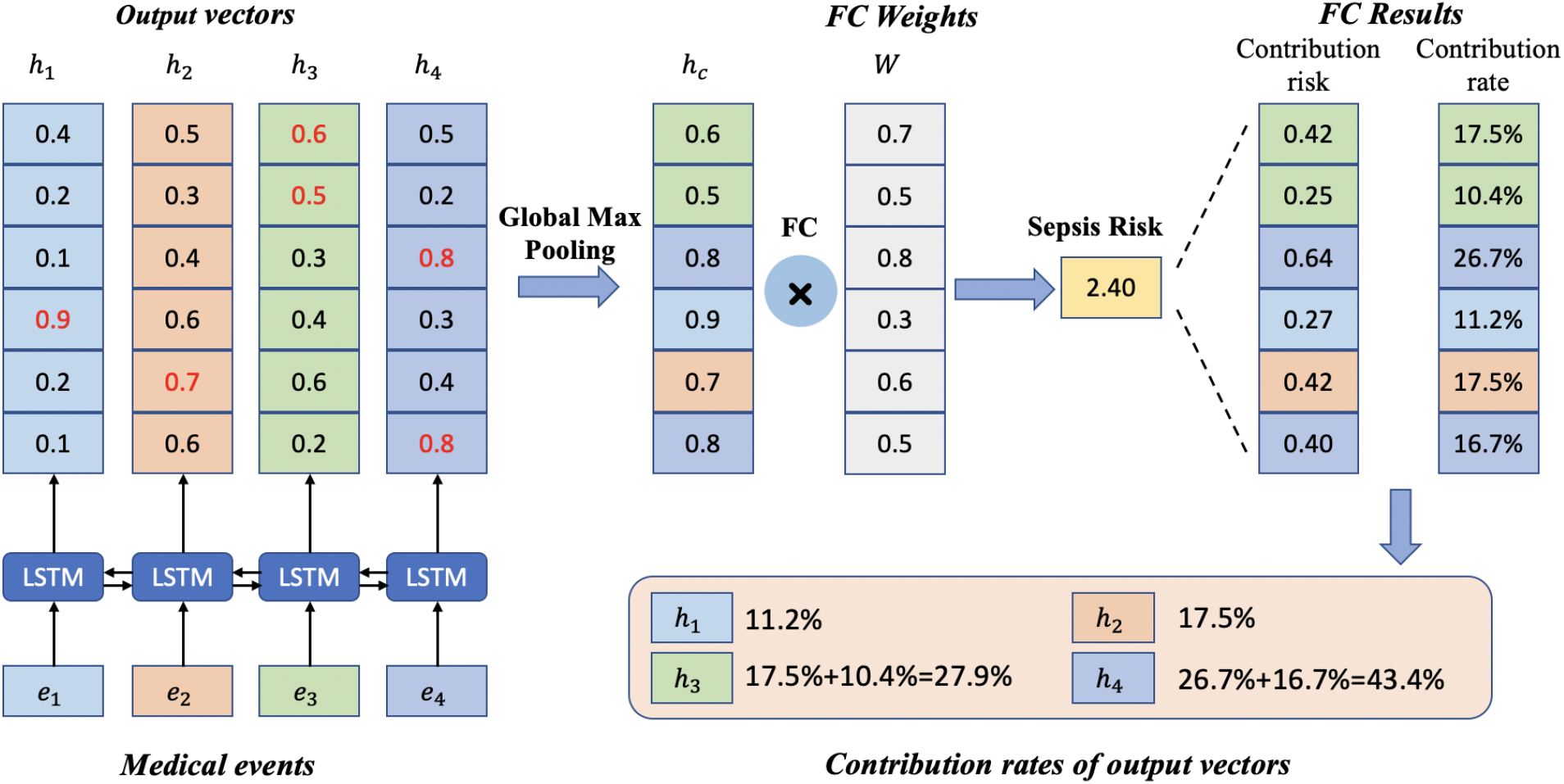
Interpretability of the proposed model with global max pooling: a toy example. Here we display 2 medical events (*e*_1_, *e*_2_) and their correpsonding output vectors (*h*_1_, *h*_2_, *h*_3_, *h*_4_). After a global max pooling layer and a fully connected layer, the model predicts the risk of sepsis onset in the next 4 hours for an individual patient. Then each output vector’s contribution is calculated by summing the corresponding dimensions’ contribution risks. Finally, the contribution of each medical event is calculated according to Eq. 5.

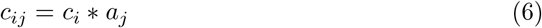

## Results and Discussion

### Experiment settings

#### Baselines

We implemented and evaluated 4 early warning scores and 3 traditional machine learning methods as baselines. The 4 early warning scores included Modified Early Warning Score (MEWS) [28], National Early Warning Score (NEWS) [29], Systemic Inflammatory Response Syndrome (SIRS) [30], and quick Sequential (Sepsis-Related) Organ Failure Assessment (qSOFA) [4]. For traditional machine learning methods, we considered logistic regression, random forest, and gradient boosting trees. Because these standard machine learning methods cannot work directly with multivariate time-series sequences, the element-wise aggregation (i.e. count, mean value, minimum value, maximum value, and standard deviation of events) of temporal features are used as model inputs.

#### Implementation details

The 4 early warning scores are calculated based on the worst value for each physiological variable within the past 24 hours before *T*_*onset*_ −4 (i.e., the last observed time point). Logistic regression and random forest are implemented with scikit-learn toolkit [31]. We implement gradient boosting trees using LightGBM [32]. For the proposed LSTM-based model, we use PyTorch [33] and the number of timesteps for LSTM is set to 100. For evaluation, 80% of the data are used for training, and 10% for validation, 10% for testing. For binary classification tasks, AUC score is used as the evaluation metric. The competition was hosted on Amazon Web Services (AWS) and experiments were conducted on a limited secure server to protect data privacy. GPUs are available to accelerate computing.

### Results

In this section, we report the performance of our proposed model on the sepsis prediction task. The results are shown in Table 2. To validate the contribution of event embeddings, time encodings, and global max pooling, we adopt ablation study and the results are shown in Table 3.

**Table 2.** AUC scores of sepsis onset prediction task on the training dataset.

| Method | Case1 | Case2 | Average |
| --- | --- | --- | --- |
| MEWS | 0.54 | 0.72 | 0.63 |
| NEWS | 0.52 | 0.72 | 0.62 |
| SIRS | 0.56 | 0.69 | 0.62 |
| qSOFA | 0.53 | 0.65 | 0.59 |
| Logistic regression | 0.89 | 0.79 | 0.84 |
| Random forest | 0.90 | 0.81 | 0.85 |
| LightGBM | 0.91 | 0.81 | 0.86 |
| Proposed model | **0.94** | **0.84** | **0.89** |

**Table 3.** Ablation study of different components (i.e. event embeddings, time encodings, and global max pooling).

| Model | AUC |
| --- | --- |
| Proposed model w/o event embeddings | 0.83 |
| Proposed model w/o time encodings | 0.89 |
| Proposed model w/o global max pooling | 0.91 |
| Proposed model | **0.94** |

#### Classification results

Table 2 summarizes the performance of various models for sepsis onset prediction on the training set. From Table 2, our model outperforms baseline models. The main reasons why our model works better are two-fold:

(1) Our model can automatically learn better patient representations as the network grows deeper and yield more accurate predictions with sufficient data. (2) Our LSTM-based model can better capture temporal information, while logistic regression, random forest, and LightGBM simply aggregate time-series features and hence suffer from information loss.

We found that machine learning-based algorithms outperformed early warning scores on both 2 cases. All 3 machine learning methods achieved similar performance on both Case 1 and Case 2. MEWS and NEWS were shown to perform better than SIRS and qSOFA on Case 2. However, the result suggested little discrimination of 4 scores on Case 1 with low AUC scores.

On the private test dataset, our proposed model achieved AUC scores of 0.940 and 0.845 for 2 use cases respectively. The official score is (0.940 + 0.845)*/*2 = 0.892.

#### Ablation study

In order to measure the effectiveness of different components (i.e. event embeddings, time encodings, and global max pooling), we adopt an ablation study to gain a better understanding of the proposed model by removing one component each time. The results of ablation study on Case 1 sepsis onset prediction are reported in Table 3. Based on the results from Table 3, the most influential component is event embeddings. By removing event embeddings, AUC score decreases 0.11. By handling irregular time intervals using time encoding, the model performance increases from 0.89 to 0.94. Besides, incorporating global max pooling causes an AUC score increase of 0.03.

### Interpretability

Generally, linear models and tree-based models can be easily interpreted because of their intuitive way of predicting output from inputs, but these models are quite simple. Although deep learning models can usually yield more accurate predictions, they usually operate as black boxes and make it unclear why the models make specific predictions. However, due to the attention mechanism and global max pooling operation, our deep learning model is interpretable. At patient level, we are able to calculate the contribution rate of each medical event for sepsis risk according to Eq. 5. Medical events with higher contribution rates contribute the most to the clinical outcome (i.e. sepsis onset in the next 4 hours).

While patient-level interpretation reveals medical events that are most influential to sepsis onset for an individual patient, population-level analysis is needed to determine the most influential medical events as well as clinical features over the entire EHR dataset. Therefore, to better understand the model’s behavior, we try to interpret our model at population-level in two ways (i.e., medical event importance, influential clinical features).

#### Medical event importance

As we can calculate the contribution rate of each medical event of each patient, we can compute each medical event’s importance on population level. For each medical event, event importance is calculated by averaging it’s contribution rates for all patients, whose EHR data contain this event.

Figure 5 shows the medical event importance (average contribution rate) over time for all patients. We can observe this plot shows an overall upward trend. This observation meets our expectation, the medical events are closer to sepsis onset are more important for our model to make predictions.

**Figure 5.**
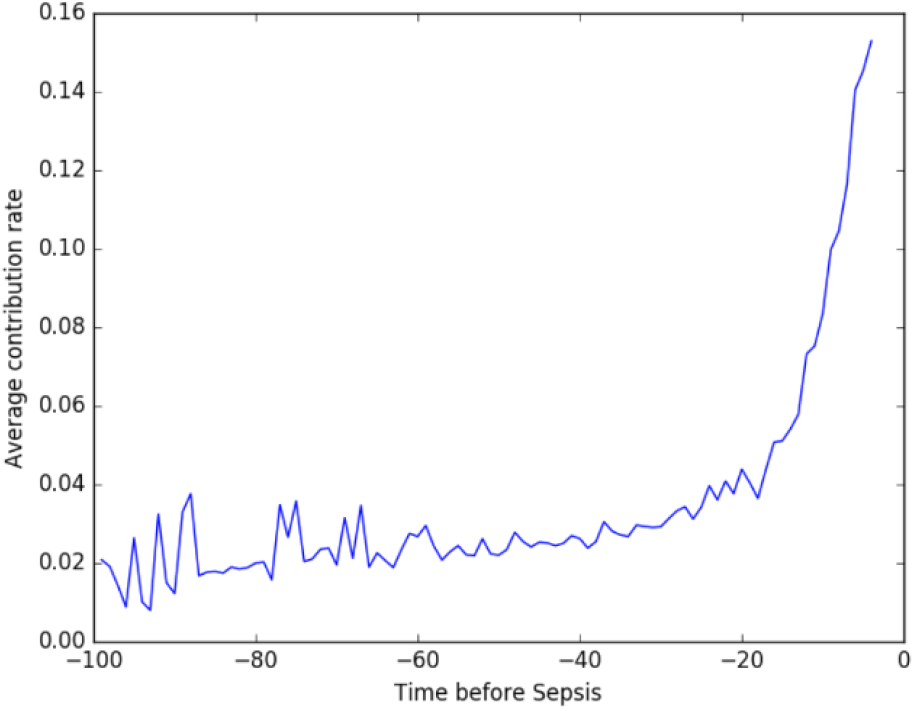
Average contribution rate of medical events over time for all patients.

**Figure 6.**
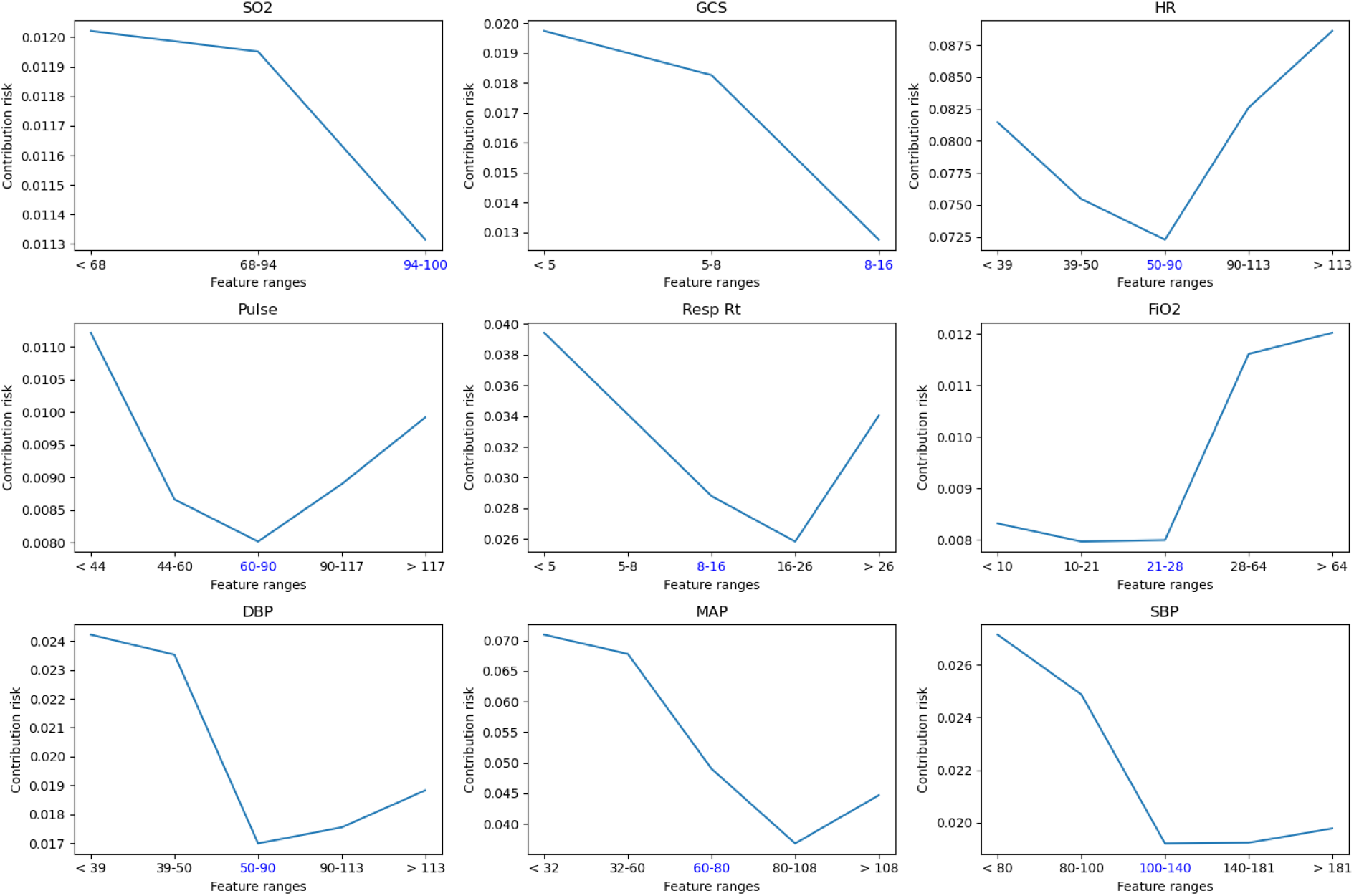
Contribution risks of top influential features found by the deep learning model. Blue-colored tick label on x-axis is the corresponding normal range of each feature.

#### Clinical feature importance

Apart from medical event importance, we also want to know which clinical features are most important for sepsis onset prediction. Similar to medical event importance, for each clinical feature, we compute its importance over all medical events on the entire population according to Eq. 6. The top influential features found by the deep learning model are shown in Figure6. The clinical features with the highest contribution to sepsis prediction are easily attainable clinical values. Thus our model suggests the development of sepsis can be predicted easily based upon items within the EHR. Interestingly, lab values traditionally associated with sepsis prediction (for example, white blood cell count and renal function) were not predictive in this model.

## Conclusion

Our team, *BuckeyeAI*, participated in the 2019 DII Challenge and ranked #2 out of 30 teams on the early prediction of sepsis onset task. In this paper, we presented our solution to sepsis onset prediction 4 hours before it occurs. For sepsis onset prediction, our proposed deep model achieved an AUC score of 0.892 and outperformed several baseline machine learning models. By incorporating event embeddings, time encodings, and global max pooling, our model yields more accurate predictions. Time encodings help handle irregular time intervals. The global pooling operation enables the model to associate the contribution of each medical event to the final clinical outcome, paving the way for interpretable clinical risk predictions.

Although we mainly focus on sepsis onset prediction in this challenge, our model is general and can be applied to other multi-variate time-series prediction problems. In addition to the superior performance, our proposed model is interpretable from an individual patient to the whole population.

## Data Availability

Cerner Health Facts is subscribed by School of Biomedical Informatics, UTHealth and a subset of data were approved for use for the 2019 DII National Data Science Challenge by Cerner and UTHealth.
Cerner Health Facts is a database that comprises de-identified EHR data from over 600 participating Cerner client hospitals and clinics in the United States and represents over 106 million unique patients. With this longitudinal, relational database-reflecting data from 2000-2016-researchers can analyze detailed sets of de-identified clinical data at the patient level. Types of data available include demographics, encounters, diagnoses, procedures, lab results, medication orders, medication administration, vital signs, microbiology, surgical cases, other clinical observations, and health systems attributes.

https://sbmi.uth.edu/dii-challenge/index-new.htm

## Acknowledgements

The authors would like to thank Dr. Lawrence Lynn for the weekly discussions of sepsis during the 2019 DII National Data Science Challenge.

## Author’s contributions

PZ conceived the project. DZ and CY developed the method. DZ conducted the experiments. DZ, CY, KB, JC, and PZ analyzed experimental results. DZ, CY, and PZ wrote the manuscript. XJ organized the 2019 DII Challenge and processed the dataset. All authors read and approved the final manuscript.

## Funding

This work was supported in part by the National Institute on Aging (R03AG064379 for KB). This project was supported in part under a grant with Lyntek Medical Technologies Inc (for DZ, CY, and PZ).

## Availability of data and materials

The source code is provided for reproducing and is available at

https://github.com/yinchangchang/DII-Challenge.

## Ethics approval and consent to participate

Not applicable.

## Consent for publication

Not applicable.

## Competing interests

Not applicable.

## Notes

### Competing Interest Statement

The authors have declared no competing interest.

### Author Declarations

Cerner Health Facts is subscribed by School of Biomedical Informatics, UTHealth and a subset of data were approved for use for the 2019 DII National Data Science Challenge by Cerner and UTHealth. Cerner Health Facts is a database that comprises de-identified EHR data from over 600 participating Cerner client hospitals and clinics in the United States and represents over 106 million unique patients. With this longitudinal, relational database-reflecting data from 2000-2016-researchers can analyze detailed sets of de-identified clinical data at the patient level. Types of data available include demographics, encounters, diagnoses, procedures, lab results, medication orders, medication administration, vital signs, microbiology, surgical cases, other clinical observations, and health systems attributes.

